## Supplementary Material for "Assessing the impact of widespread respirator use in curtailing COVID-19 transmission in the United States"

#### 1 Supplementary Material for the Basic Model

Table S1: Description of the state variables and parameters of the basic model (2.1).

| State variable | Description |
|---|---|
| $S$ | Population of susceptible individuals |
| $E$ | Population of exposed (newly-infected but not yet infectious) individuals |
| $P$ | Population of pre-symptomatically-infectious individuals |
| $I$ | Population of symptomatically-infectious individuals |
| $A$ | Population of asymptomatically-infectious individuals |
| $H$ | Population of hospitalized individuals |
| $R$ | Population of recovered individuals |
| **Parameter** | **Description** |
| $\beta$ | Overall community transmission rate |
| $\eta_P$ | Modification parameter for infectiousness of pre-symptomatic individuals |
| $\eta_I$ | Modification parameter for infectiousness of symptomatic individuals |
| $\eta_A$ | Modification parameter for infectiousness of asymptomatic individuals |
| $\eta_H$ | Modification parameter for infectiousness of hospitalized individuals |
| $c_m$ | Community face masks compliance |
| $0 < \varepsilon_m < 1$ | Protective efficacy of face masks |
| $\sigma_E$ | Progression rate of exposed individuals to the pre-symptomatic infectious stage |
| $\sigma_P$ | Progression rate from pre-symptomatic to symptomatic or asymptomatic class |
| $r$ | Proportion of pre-symptomatic individuals who become symptomatic |
| $1-r$ | Proportion of pre-symptomatic infectious individuals who become asymptomatic |
| $\phi_I$ | Hospitalization rate for symptomatically-infectious individuals |
| $\gamma_A$ | Recovery rate for asymptomatically-infectious individuals |
| $\gamma_I$ | Recovery rate for symptomatically-infectious individuals |
| $\gamma_H$ | Recovery rate for hospitalized individuals |
| $\delta_I$ | Disease-induced mortality rate for symptomatically-infectious individuals |
| $\delta_H$ | Disease-induced mortality rate for hospitalized individuals |

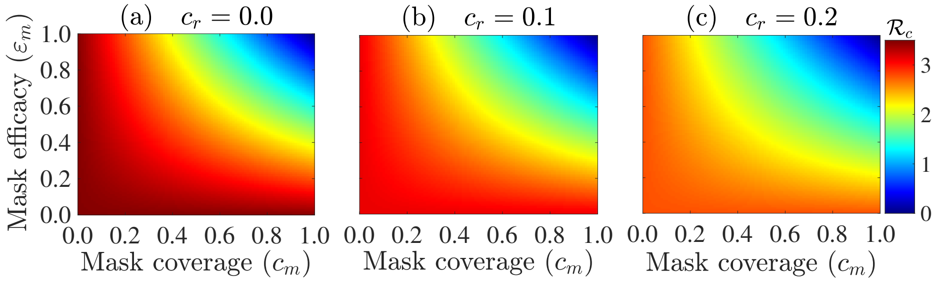


Figure S1: Heatmap of the control reproduction number ($\mathbb{R}_c$) of the basic model (2.1) in the main text, as a function of the face masks compliance ($c_m$) and efficacy ($\varepsilon_m$). (a) No reduction in community transmission from baseline due to the implementation of other NPIs (i.e., $c_r = 0$). (b) 10% reduction in community transmission due to implementation of other NPIs (i.e., $c_r = 0.1$). (c) 20% reduction in community transmission due to the implementation of other NPIs (i.e., $c_r = 0.2$). The other parameter values used in the simulations are as given in Tables 1 and 2 (a) in the main text.

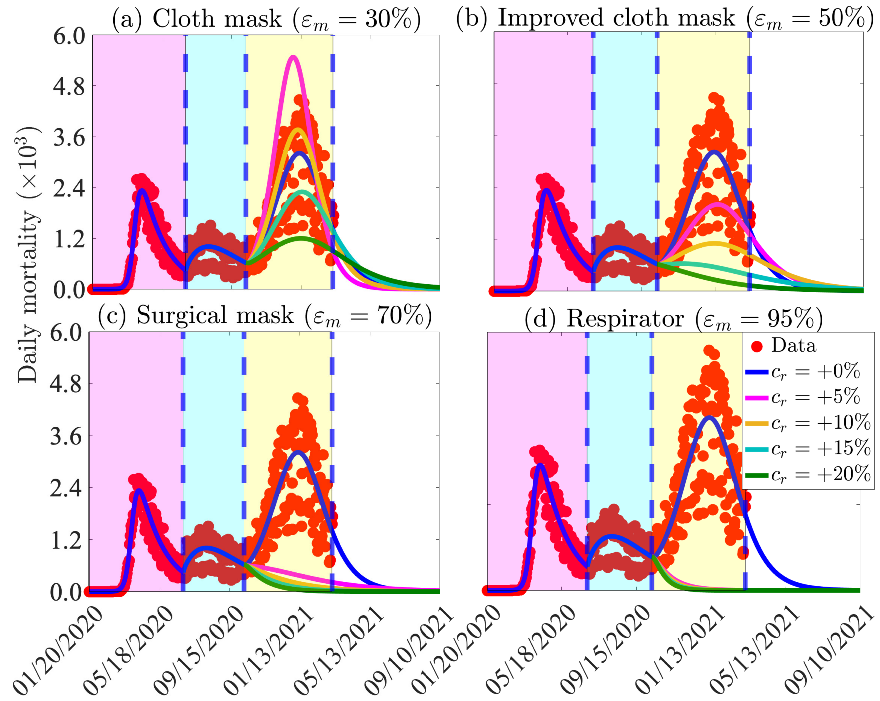


Figure S2: Simulations of the basic model (2.1) to assess the combined impact of face mask use strategy and reduction in community transmission due to other NPIs (implemented at the beginning of the third wave) on daily COVID-19 mortality in the US (a) Only cloth masks are used. (b) Only improved cloth masks are used. (c) Only surgical masks are used. (d) Only N95 respirator (or equivalent) are used. For the blue curves (baseline scenario) in Figures (a)-(d), the mask efficacy is maintained at 50% (i.e., $\varepsilon_m = 0.5$). The other parameter values used in the simulations are given in Tables 1 and 2 of the main text. The shaded magenta and green regions represent the beginning and end of the first and second pandemic waves in the US, respectively. The shaded yellow region represents the beginning of the third pandemic wave until March 11, 2021. Dashed vertical blue lines demarcate the three waves.

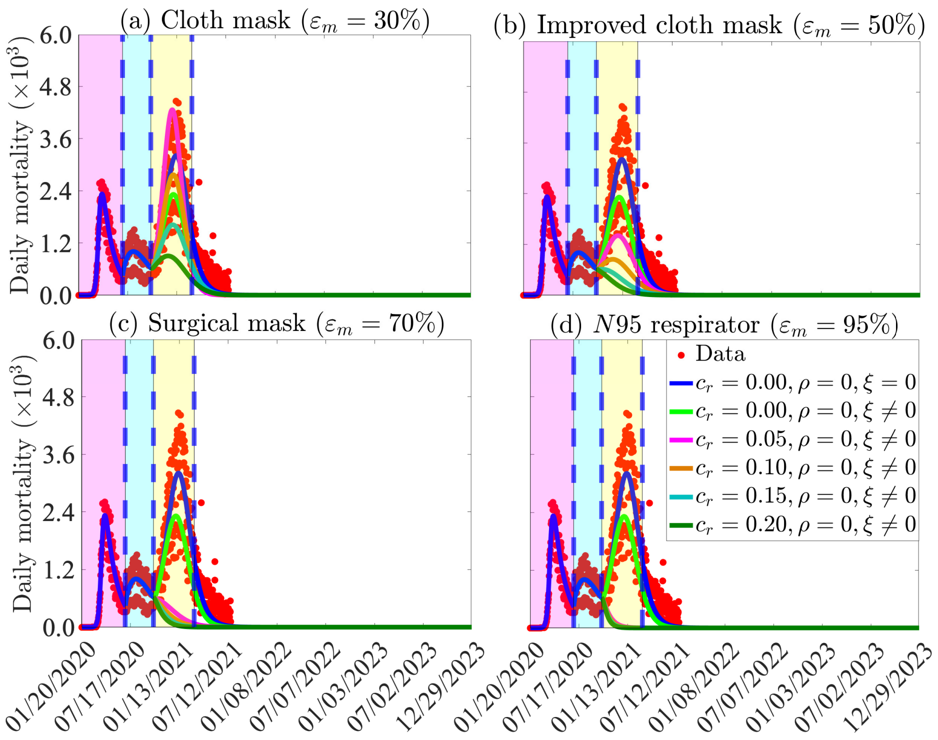


Figure S3: Simulations of the basic model (2.1) in the main text to assess the combined impact of masks, vaccination, permanent natural and vaccine-derived immunity, and reduction in community transmission due to other NPIs on the daily COVID-19 mortality as a function of time, and various mask types, for the US. Reduction in community transmission due to other NPIs and vaccination are implemented at the beginning of the third wave (i.e., from October 12, 2020). For these simulations, only the Pfizer vaccine is used (with estimated efficacy of $95\%$) and the vaccination rate is set at $\xi = 7.4 \times 10^4$ *per* day (i.e.,$250,000$ individuals are fully-vaccinated daily). (a) Only cloth masks are used. (b) Only improved cloth masks are used. (c) Only surgical masks are used. (d) Only respirators (or equivalent) are used. For the blue and light green curves (baseline scenarios) in Figures (a)-(d), the mask efficacy is maintained at $50\%$ (i.e., $\varepsilon_m = 0.5$). The other parameter values used in the simulations are given in Tables 1 and 2 of the main text. The shaded magenta and green regions represent the beginning and end of the first and second pandemic waves in the US, respectively. The shaded yellow region represents the beginning of the third pandemic wave until March 11, 2021. Dashed vertical blue lines demarcate the three waves.

### 2 Supplementary Material for the Extended Model

#### 2.1 Formulation of the Extended Model

The basic model (2.1) is extended to account for behavioural changes with respect to face masks usage by subdividing the total population into the three subgroups of those who do not face masks at all (group 1), those who only wear fabric masks (group 2) and those who only wear N-95 respirators or equivalent (group 3). We further introduce the new parameters $\alpha_{ij}$, with $i, j = 1, 2, 3$ and $i \neq j$, for the back-and-forth transitions between the three groups.

| Mask use behavior change parameter | Description |
|---|---|
| $\alpha_{12}$ | Transition rate from group 1 to group 2 |
| $\alpha_{13}$ | Transition rate from group 1 to group 3 |
| $\alpha_{21}$ | Transition rate from group 2 to group 1 |
| $\alpha_{31}$ | Transition rate from group 3 to group 1 |
| $\alpha_{23}$ | Transition rate from group 2 to group 3 |
| $\alpha_{32}$ | Transition rate from group 3 to group 2 |

Table S2: Change of behavior parameters, $\alpha_{ij}; i = 1, 2, 3(i \neq j)$, for the extended model (S.1)-(S.3).

The equations for the rate of change of the state variables of the extended multigroup model are given below.

##### 2.1.1 Equations for Group 1 dynamics: individuals who do not wear masks in public

$$\begin{aligned}
\dot{S}_1 &= \alpha_{21}S_2 + \alpha_{31}S_3 - (\lambda_1 + \alpha_{12} + \alpha_{13})S_1, \\
\dot{E}_1 &= \alpha_{21}E_2 + \alpha_{31}E_3 + \lambda_1 S_1 - (\alpha_{12} + \alpha_{13} + \sigma_E)E_1, \\
\dot{P}_1 &= \alpha_{21}P_2 + \alpha_{31}P_3 + \sigma_E E_1 - (\alpha_{12} + \alpha_{13} + \sigma_P)P_1, \\
\dot{I}_1 &= \alpha_{21}I_2 + \alpha_{31}I_3 + r\sigma_P P_1 - (\alpha_{12} + \alpha_{13} + \phi_I + \gamma_I + \delta_I)I_1, \\
\dot{A}_1 &= \alpha_{21}A_2 + \alpha_{31}A_3 + (1-r)\sigma_P P_1 - (\alpha_{12} + \alpha_{13} + \gamma_A)A_1, \\
\dot{H}_1 &= \alpha_{21}H_2 + \alpha_{31}H_3 + \phi_I I_1 - (\alpha_{12} + \alpha_{13} + \gamma_H + \delta_H)H_1, \\
\dot{R}_1 &= \alpha_{21}R_2 + \alpha_{31}R_3 + \gamma_I I_1 + \gamma_A A_1 + \gamma_H H_1 - (\alpha_{12} + \alpha_{13})R_1,
\end{aligned} \tag{S.1}$$

where $\lambda_1$ is the *force of infection*, defined by:

$$\begin{aligned}
\lambda_1 &= \beta_1 \frac{(\eta_P P_1 + \eta_I I_1 + \eta_A A_1 + \eta_H H_1)}{N} \\
&+ (1 - c_{m2}\varepsilon_{o2})\beta_2 \frac{(\eta_P P_2 + \eta_I I_2 + \eta_A A_2 + \eta_H H_2)}{N} \\
&+ (1 - c_{m3}\varepsilon_{o3})\beta_3 \frac{(\beta_P P_3 + \eta_I I_3 + \eta_A A_3 + \eta_H H_3)}{N},
\end{aligned}$$

where $0 < \varepsilon_{o2} < 1$ and $0 < \varepsilon_{o3} < 1$ represent the outward protective efficacies of fabric and N-95 masks, respectively, to prevent the transmission of infection to a susceptible individual who does not wear a face mask in public, while $0 < c_{m2} < 1$ and $0 < c_{m3} < 1$, are the face mask compliance defined as the proportion of individuals who upon switching their behavior to wearing masks, wear the masks habitually in public.

##### 2.1.2 Equations for Group 2 dynamics: individuals who wear cloths and surgical masks only

$$\begin{aligned}
\dot{S}_2 &= \alpha_{12}S_1 + \alpha_{32}S_3 - (\lambda_2 + \alpha_{21} + \alpha_{23})S_2, \\
\dot{E}_2 &= \lambda_2 S_2 + \alpha_{12}E_1 + \alpha_{32}E_3 - (\sigma_E + \alpha_{21} + \alpha_{23})E_2, \\
\dot{P}_2 &= \sigma_E E_2 + \alpha_{12}P_1 + \alpha_{32}P_3 - (\sigma_P + \alpha_{21} + \alpha_{23})P_2, \\
\dot{I}_2 &= r\sigma_P P_2 + \alpha_{12}I_1 + \alpha_{32}I_3 - (\phi_I + \gamma_I + \delta_I + \alpha_{21} + \alpha_{23})I_2, \\
\dot{A}_2 &= (1-r)\sigma_P P_2 + \alpha_{12}A_1 + \alpha_{32}A_3 - (\gamma_A + \alpha_{21} + \alpha_{23})A_2, \\
\dot{H}_2 &= \phi_I I_2 + \alpha_{12}H_1 + \alpha_{32}H_3 - (\gamma_H + \delta_H + \alpha_{21} + \alpha_{23})H_2, \\
\dot{R}_2 &= \gamma_I I_2 + \gamma_A A_2 + \gamma_H H_2 + \alpha_{12}R_1 + \alpha_{32}R_3 - (\alpha_{21} + \alpha_{23})R_2,
\end{aligned} \tag{S.2}$$

where,

$$
\begin{aligned}
\lambda_2 &= (1 - c_{m2}\varepsilon_{i2})\beta_1 \frac{(\eta_P P_1 + \eta_I I_1 + \eta_A A_1 + \eta_H H_1)}{N} \\
&+ (1 - c_{m2}\varepsilon_{i2})(1 - c_{m2}\varepsilon_{o2})\beta_2 \frac{(\eta_P P_2 + \eta_I I_2 + \eta_A A_2 + \eta_H H_2)}{N} \\
&+ (1 - c_{m2}\varepsilon_{i2})(1 - c_{m3}\varepsilon_{o3})\beta_3 \frac{(\eta_P P_3 + \eta_I I_3 + \eta_A A_3 + \eta_H H_3)}{N}.
\end{aligned}
$$

The parameters $0 < \varepsilon_{o2} < 1$ and $0 < \varepsilon_{o3} < 1$ represent the outward protective efficacies of fabric and N-95 masks, respectively, to prevent the transmission of infection to a susceptible individual who wears a cloth face mask in public, and $0 < \varepsilon_{i2} < 1$ represents the inward protective efficacy of fabric face masks to prevent the acquisition of infection from an infectious individual.

###### 2.1.3 Equations for Group 3 dynamics: individuals who wear N95 respirators (or equivalent) only

$$
\begin{aligned}
\dot{S}_3 &= \alpha_{13}S_1 + \alpha_{23}S_2 - (\lambda_3 + \alpha_{31} + \alpha_{32})S_3, \\
\dot{E}_3 &= \lambda_3 S_3 + \alpha_{13}E_1 + \alpha_{23}E_2 - (\sigma_E + \alpha_{31} + \alpha_{32})E_3, \\
\dot{P}_3 &= \sigma_E E_3 + \alpha_{13}P_1 + \alpha_{23}P_2 - (\sigma_P + \alpha_{31} + \alpha_{32})P_3, \\
\dot{I}_3 &= r\sigma_P P_3 + \alpha_{13}I_1 + \alpha_{23}I_2 - (\phi_I + \gamma_I + \delta_I + \alpha_{31} + \alpha_{32})I_3, \\
\dot{A}_3 &= (1 - r)\sigma_P P_3 + \alpha_{13}A_1 + \alpha_{23}A_2 - (\gamma_A + \alpha_{31} + \alpha_{32})A_3, \\
\dot{H}_3 &= \phi_I I_3 + \alpha_{13}H_1 + \alpha_{23}H_2 - (\gamma_H + \delta_H + \alpha_{31} + \alpha_{32})H_3, \\
\dot{R}_3 &= \gamma_I I_3 + \gamma_A A_3 + \gamma_H H_3 + \alpha_{13}R_1 + \alpha_{23}R_2 - (\alpha_{31} + \alpha_{32})R_3,
\end{aligned}
\tag{S.3}
$$

where,

$$
\begin{aligned}
\lambda_3 &= (1 - c_{m3}\varepsilon_{i3})\beta_1 \frac{(\eta_P P_1 + \eta_I I_1 + \eta_A A_1 + \eta_H H_1)}{N} \\
&+ (1 - c_{m3}\varepsilon_{i3})(1 - c_{m2}\varepsilon_{o2})\beta_2 \frac{(\eta_P P_2 + \eta_I I_2 + \eta_A A_2 + \eta_H H_2)}{N} \\
&+ (1 - c_{m3}\varepsilon_{i3})(1 - c_{m3}\varepsilon_{o3})\beta_3 \frac{(\eta_P P_3 + \eta_I I_3 + \eta_A A_3 + \eta_H H_3)}{N}.
\end{aligned}
$$

Table S3: Description of the state variables of the extended model (S.1) − (S.3).

| Variable | Description |
|---|---|
| $S_1$ | Population of susceptible individuals who do not wear face masks |
| $S_2$ | Population of susceptible individuals who wear fabric (cloth or surgical) face masks |
| $S_3$ | Population of susceptible individuals who wear N-95 respirators |
| $E_1$ | Population of exposed (newly-infected) individuals who do not wear face masks |
| $E_2$ | Population of exposed (newly-infected) individuals who wear fabric face masks |
| $E_3$ | Population of exposed (newly-infected) individuals who wear N-95 respirators |
| $P_1$ | Population of pre-symptomatic infectious individuals who do not wear face masks |
| $P_2$ | Population of pre-symptomatic infectious individuals who wear fabric face masks |
| $P_3$ | Population of pre-symptomatic infectious individuals who wear N-95 respirators |
| $I_1$ | Population of symptomatically-infectious individuals who do not wear face masks |
| $I_2$ | Population of symptomatically-infectious individuals who wear fabric face masks |
| $I_3$ | Population of symptomatically-infectious individuals who wear N-95 respirators |
| $A_1$ | Population of asymptomatically-infectious individuals who do not wear face masks |
| $A_2$ | Population of asymptomatically-infectious individuals who wear fabric face masks |
| $A_2$ | Population of asymptomatically-infectious individuals who wear N-95 respirators |
| $H_1$ | Population of hospitalized individuals who do not wear face masks |
| $H_2$ | Population of hospitalized individuals who wear fabric face masks |
| $H_3$ | Population of hospitalized individuals who wear N-95 respirators |
| $R_1$ | Population of recovered individuals who do not wear face masks |
| $R_2$ | Population of recovered individuals who wear fabric face masks |
| $R_3$ | Population of recovered individuals who wear N-95 respirators |

Table S4: Description of the parameters of the extended model (S.1)-(S.3)

| **Parameters** | **Description** |
|---|---|
| $\beta_1$ | Effective contact rate for individuals who do not wear face masks |
| $\beta_2(\beta_3)$ | Effective contact rate for individuals who wear fabric (N-95) face masks |
| $\epsilon_{o2}(\epsilon_{i2})$ | Outward (inward) efficacy of fabric face masks |
| $\epsilon_{o3}(\epsilon_{i3})$ | Outward (inward) efficacy of N-95 respirators |
| $c_{m2}(c_{m3})$ | Fabric/surgical (N-95) mask compliance (i.e., the proportion of individuals in the community who wear the appropriate type of masks consistently) |
| $\alpha_{12}(\alpha_{13})$ | Rate at which individuals who do not wear masks habitually choose to become habitual fabric (N-95) mask wearers |
| $\alpha_{21}(\alpha_{31})$ | Rate at which individuals who wear fabric (N-95) masks habitually choose to stop wearing masks |
| $\alpha_{23}(\alpha_{32})$ | Rate at which individuals who wear fabric (N-95) masks habitually switch to become habitual N-95 (fabric) mask wearers |
| $\sigma_E$ | Rate at which exposed individuals progress to the pre-symptomatic infectious stage |
| $\sigma_P$ | Rate at which pre-symptomatic infectious individuals progress to symptomatically-infectious or asymptomatically-infectious stage |
| $r$ | Proportion of pre-symptomatic infectious individuals who become symptomatically-infectious |
| $\phi_I$ | Hospitalization rate for symptomatically-infectious individuals |
| $\gamma_A$ | Recovery rate for asymptomatically-infectious individuals |
| $\gamma_I$ | Recovery rate for symptomatically-infectious individuals |
| $\gamma_H$ | Recovery rate for hospitalized individuals |
| $\delta_I$ | Disease-induced mortality rate for symptomatically-infectious individuals |
| $\delta_H$ | Disease-induced mortality rate for hospitalized individuals |

#### 2.2 Data Fitting and Parameter Estimation of the Extended Model

As with the single group model, a least squares approach is used to fit the extended model to cumulative mortality data for the US. This process leads to the estimation of 14 of the model parameters. The estimated parameters and their 95% confidence intervals are presented in Tables S5-S6, while the plot of the model fit together with the plot of the data and output of the model (obtained using the fitted parameters and other fixed parameters) are presented in Figure S4.

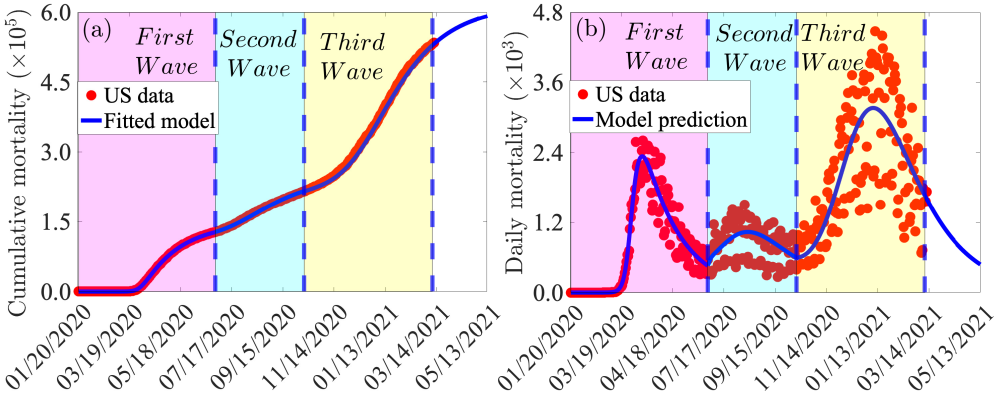


Figure S4: (a) Time series plot of the actual cumulative mortality data for the US (red dots) and predicted cumulative mortality for the US from the model (S.1)-(S.3). (b) Simulations of the model (S.1)-(S.3) using the estimated parameters in Tables S5-S6 and the fixed parameters in Table 2 of the main text illustrating the three COVID-19 waves observed in the US during the periods from January 20, 2020 to June 30, 2020 (first pandemic wave), July 1, 2020 to October 11, 2020 (second pandemic wave), and October 12, 2020 to March 11, 2021 (third pandemic wave). Dashed vertical blue lines demarcate the pandemic waves.

Table S5: Estimated (fitted) baseline parameter values and confidence intervals (CIs) for the basic model (S.1)-(S.3) using COVID-19 mortality data for the U.S. for the period from (a) January 20, 2020 to June 30, 2020 (first pandemic wave), (b) July 1, 2020 to October 11, 2020 (second pandemic wave), and (c) October 12, 2020 to March 11, 2021 (third pandemic wave). In addition to using the fixed parameters in Table 2 of the main text, we set $e_{i2} = 0.70, e_{o2} = 0.60, e_{i3} = 0.95, e_{o3} = 0.85$, and $c_{m3} = 1.00$ in these simulations. Further, we set $c_{m2} = 0.50$, for the first wave and $c_{m2} = 0.60$ for the second wave. The unit of each of the rate parameters is *per* day.

| | (a) First wave (January 20-June 30, 2020) | | (b) Second wave (July 1-October 11, 2020) | |
|---|---|---|---|---|
| **Parameter** | **Estimated value** | **95% confidence interval** | **Estimated value** | **95% confidence interval** |
| $\beta_1$ | 0.96122737 | $[0.71916052, 1.46785952]$ | 0.96122737 | $[0.71916052, 1.46785952]$ |
| $\beta_2$ | 0.10115487 | $[0.00073967, 0.20432930]$ | 0.10115487 | $[0.00073967, 0.20432930]$ |
| $\beta_3$ | 0.07030497 | $[0.00054567, 0.13471250]$ | 0.07030497 | $[0.00054567, 0.13471250]$ |
| $\alpha_{12}$ | 0.00077134 | $[0.00036180, 0.00098147]$ | 0.00077134 | $[0.00036180, 0.00098147]$ |
| $\alpha_{13}$ | 0.00024978 | $[0.00010609, 0.00039998]$ | 0.00024978 | $[0.00010609, 0.00039998]$ |
| $\alpha_{23}$ | 0.00000027 | $[0.00000010, 0.00000056]$ | 0.00000027 | $[0.00000010, 0.00000056]$ |
| $\alpha_{21}$ | 0.00000041 | $[0.00000000, 0.00000218]$ | 0.00000041 | $[0.00000000, 0.00000218]$ |
| $\alpha_{31}$ | 0.00000007 | $[0.00000000, 0.00000055]$ | 0.00000007 | $[0.00000000, 0.00000055]$ |
| $\alpha_{32}$ | 0.00000014 | $[0.00000000, 0.00000059]$ | 0.00000014 | $[0.00000000, 0.00000059]$ |
| $\gamma_I$ | 0.10104010 | $[0.01000468, 0.17032509]$ | 0.10104010 | $[0.01000468, 0.17032509]$ |
| $\gamma_A$ | 0.29170892 | $[0.23812198, 0.36529185]$ | 0.29170892 | $[0.23812198, 0.36529185]$ |
| $\gamma_H$ | 0.02324779 | $[0.02261236, 0.02396185]$ | 0.02324779 | $[0.02261236, 0.02396185]$ |
| $\delta_I$ | 0.00001204 | $[0.00000000, 0.00003358]$ | 0.00001204 | $[0.00000000, 0.00003358]$ |
| $\delta_H$ | 0.00008315 | $[0.00005258, 0.00010324]$ | 0.00008315 | $[0.00005258, 0.00010324]$ |

Table S6: Estimated (fitted) baseline parameter values and the 95% confidence intervals (CIs) for the model (S.1)-(S.3) using COVID-19 mortality data for the US for the period from October 12, 2020 to March 11, 2021 (third pandemic wave). In addition to the fixed parameters in Table 2 of the main text, we set $c_{m2} = 0.70, e_{i2} = 0.70, e_{o2} = 0.60, c_{m3} = 1.00, e_{i3} = 0.95$, and $e_{o3} = 0.85$ in these simulations. The unit of each of the rate parameters is *per* day.

| **Parameter** | **Estimated value** | 95% **confidence interval** |
|---|---|---|
| $\beta_1$ | 1.10458793 | [0.82856750, 1.40788344] |
| $\beta_2$ | 0.06474221 | [0.04688231, 0.08983824] |
| $\beta_3$ | 0.01508329 | [0.01847386, 0.02569115] |
| $\alpha_{12}$ | 0.00141070 | [0.00100000, 0.00186259] |
| $\alpha_{13}$ | 0.00107707 | [0.00100000, 0.00159978] |
| $\alpha_{23}$ | 0.00000050 | [0.00000000, 0.00000336] |
| $\alpha_{21}$ | 0.00004672 | [0.00002135, 0.00007750] |
| $\alpha_{31}$ | 0.00000465 | [0.00000364, 0.00000885] |
| $\alpha_{32}$ | 0.00000425 | [0.00000211, 0.00000723] |
| $\gamma_I$ | 0.18722730 | [0.08600002, 0.25622129] |
| $\gamma_A$ | 0.37636579 | [0.27600001, 0.47844102] |
| $\gamma_H$ | 0.02380258 | [0.01199710, 0.05552252] |
| $\delta_I$ | 0.00011685 | [0.00010000, 0.00061358] |
| $\delta_H$ | 0.00159489 | [0.00112881, 0.00195805] |

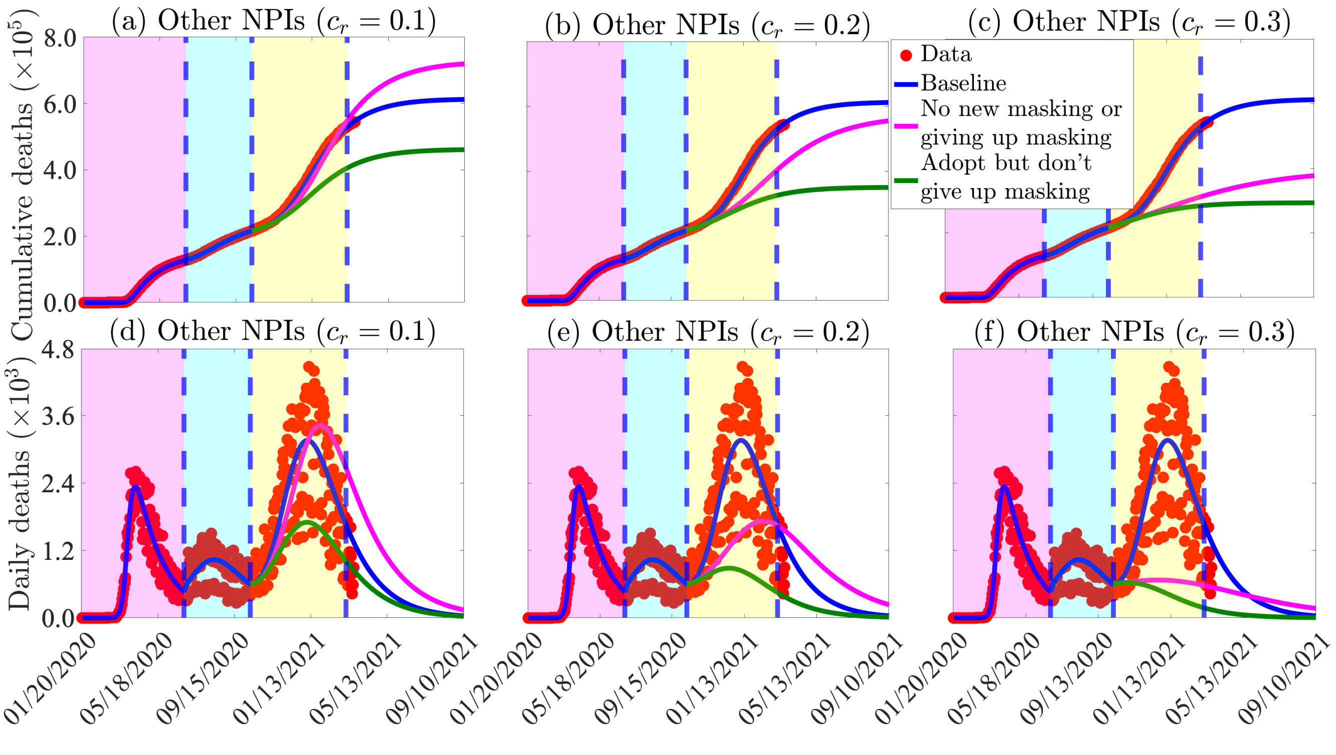


Figure S5: Simulations of the extended model (S.1)-(S.3). Figures (a)-(c): Assessment of the impact of increases in other NPIs that reduce community transmission on cumulative mortality. (a) $c_r = 0.1$. (b) $c_r = 0.2$. (c) $c_r = 0.3$. The values of the other parameters of the extended model are as given in Table 1 of the main text and Table S5-S6. The shaded magenta and green regions represent the beginning and end of the first and second pandemic waves in the U.S., respectively. The shaded yellow region represent the beginning of the third pandemic wave until March 11, 2021. Dashed vertical blue lines demarcate the three waves.

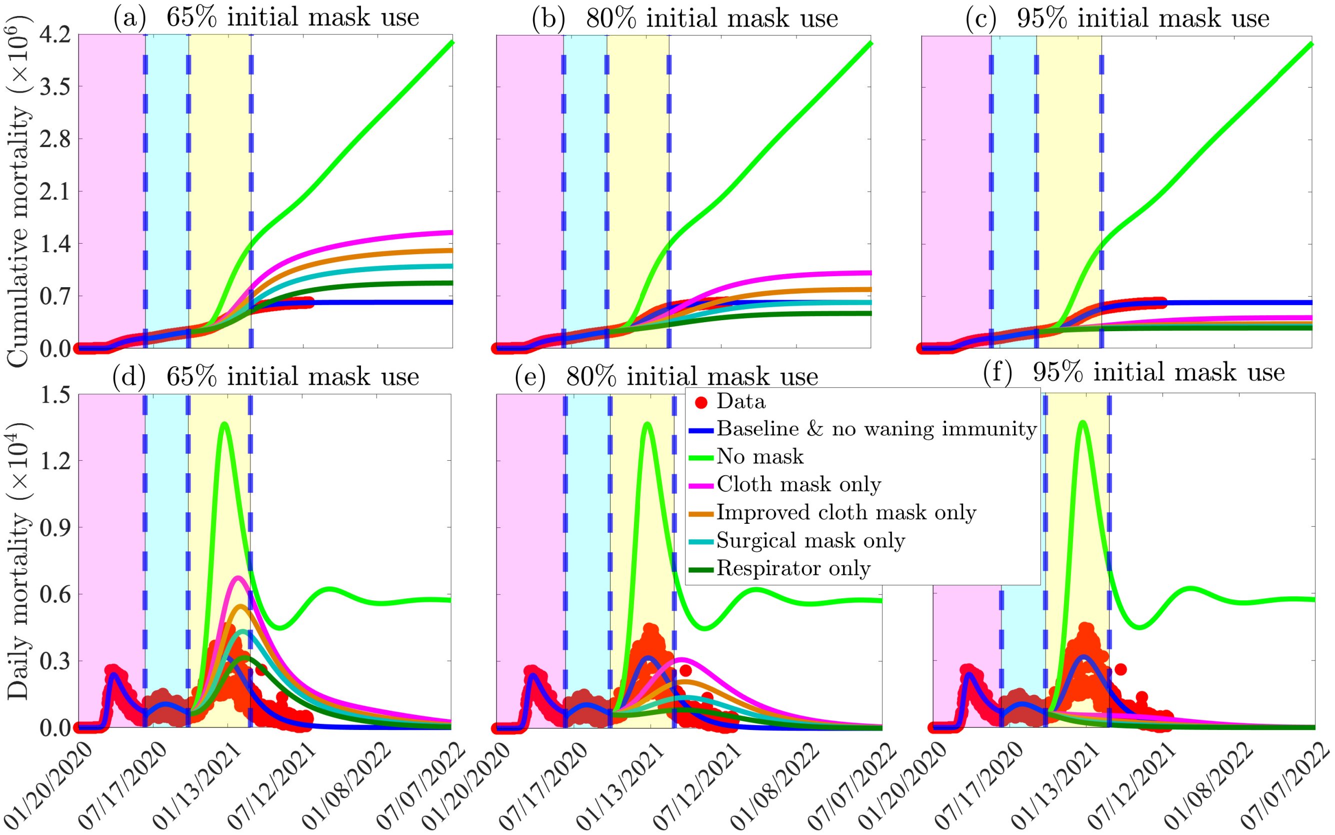


Figure S6: Simulations of the extended model (S.1)-(S.3) to assess the combined impact of the total initial size of mask wearers, the mask type (or quality), and waning natural immunity on (a)-(c) the cumulative COVID-19 mortality and (d)-(f) the daily COVID-19 mortality in the US. (a) and (d): total initial size of mask wearers is 65%; (b) and (e): total initial size of mask wearers is 80%; (c) and (f): total initial size of mask wearers is 95%. It is assumed that the average duration of natural immunity is six months, which corresponds to $\rho = 5.6 \times 10^3$ *per* day. The values of the other parameters of the extended model used in the simulations are as given in Table 1 (of the main text) and Tables S5-S6 in the Supplementary Material. The shaded magenta and green regions represent the beginning and end of the first and second pandemic waves in the US, respectively. The shaded yellow region represents the beginning of the third pandemic wave until March 11, 2021. Dashed vertical blue lines demarcate the three waves.

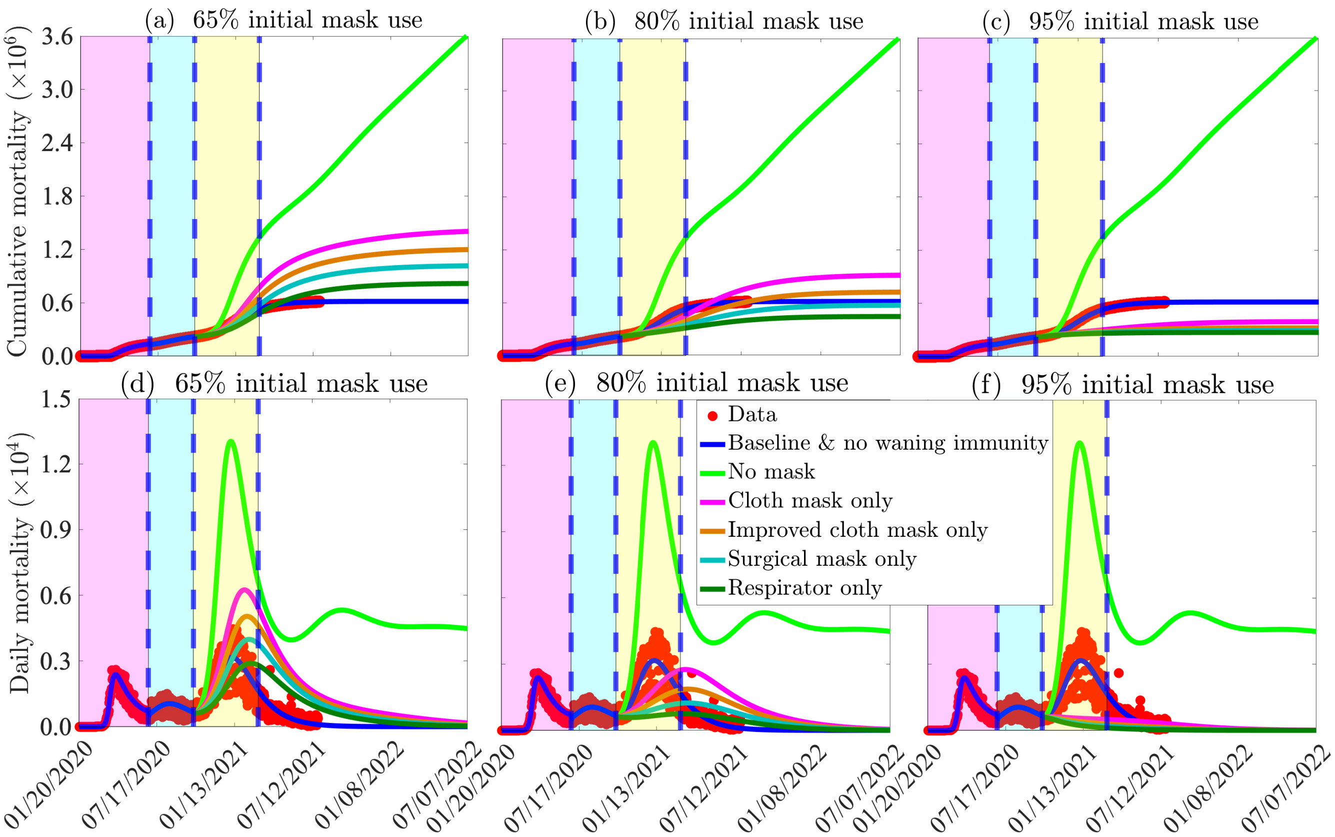


Figure S7: Simulations of the extended model (S.1)-(S.3) to assess the combined impact of the total initial size of mask wearers, the mask type (or quality), waning natural immunity, and vaccination implemented at the beginning of the third wave (i.e., from October 12, 2020) on (a)-(c) the cumulative COVID-19 mortality and (d)-(f) the daily COVID-19 mortality in the US. (a) and (d): total initial size of mask wearers is 65%; (b) and (e): total initial size of mask wearers is 80%; (c) and (f): total initial size of mask wearers is 95%. For these simulations, only the Pfizer vaccine is used (with estimated efficacy of 95%) and the vaccination rate is set at $\xi = 7.4 \times 10^4$ *per* day (i.e.,$250,000$ individuals are fully-vaccinated daily). The vaccine-derived and natural immunity wane after six months (which corresponds to $\rho = 5.6 \times 10^3$ *per* day). The values of the other parameters of the extended model used in the simulations are as given in Table 1 (of the main text) and Tables S5-S6 in the Supplementary Material. The shaded magenta and green regions represent the beginning and end of the first and second pandemic waves in the US, respectively. The shaded yellow region represents the beginning of the third pandemic wave until March 11, 2021. Dashed vertical blue lines demarcate the three waves.
